# The cost of sponsored research at institutions of higher learning in low- and middle-income countries: a cross-sectional study

**DOI:** 10.1101/2024.05.02.24306751

**Authors:** Ahaz T. Kulanga, Gibson E. Kapanda, John A. Bartlett, Charles Muiruri, Stef de Jong

## Abstract

**Introduction:** Research in low- and middle-income countries (LMICs) is challenged with poor investment by both in-country governments and philanthropies. Although research grants cover costs directly associated with specific research projects, they do not fully cover costs that are indirectly attributable to specific projects. The relationship between grant funding and actual cost of paying for such projects at institutions of higher learning (IHL) in LMICs is poorly understood. Our purpose was to evaluate unaccounted costs associated with grants and explore the system level factors that support or hinder the goal of “breaking even”.

**Materials and methods:** We used a descriptive retrospective cross-sectional design to evaluate unaccounted costs and qualitative interviews with stakeholders at three prominent medical universities in Tanzania. Data were obtained from annual audited Final Reports and Final Financial Reports for biomedical and social and behavioral sciences (non-biomedical) sponsored projects funded from 2007 through 2017.

**Findings:** A total of 17 projects were included in the study, of which 6 (35.3%) were biomedical and 11 (64.7%) non-biomedical. The median total amount of project funding for all biomedical research projects was US$ 544,084; interquartile range (IQR) [89,268-1,226,570]. These projects had median unaccounted costs of US$ 186,403 (IQR) [30,583– 420,223]. The median funding for non-biomedical sponsored research projects was US$ 902,999 (IQR) [468,259–1,951,212] and unaccounted costs were US$ 112,875 (IQR) [58,532–243,902]. 27 faculty and staff at study sites participated in interviews. Three themes emerged-costing practices, unaccounted-for costs, and budget negotiating power.

**Conclusion:** The study demonstrated that there was a substantial amount of unaccounted-for costs for sponsored projects. Costing and financial practices associated with sponsored research projects were weak, coupled with lopsided negotiating power with funding agencies. Funding agencies and institutions in this study should work to reduce the inequity in research costs.

## Introduction

To compete successfully for external support, institutions of higher learning (IHLs) must invest in the research environment including competent investigators, laboratories, and data systems among others and demonstrate their proficiencies. With these investments, IHLs reap economic and academic prestige benefits demonstrated by academic rankings which are closely associated with institutions’ research capabilities^1^.

In high-income countries (HIC), governments and philanthropies are key external funders of research in IHLs. With continual growth of economies in low- and middle-income countries (LMICs) such as African countries, there has been an impetus to recognize the value of research for sustainable development. As a result, in 2007 African leaders pledged to intensify government funding of research infrastructure^2^. Regrettably, this pledge has not been realized—most research in LMICs is funded by international organizations. Governments commit minimal or no portion of their budgets to research. For example, Franzen et al.^3^ observed that research in LMICs is challenged with poor investment by both governments and in-country philanthropies. It is estimated that almost 80% of research funding in LMICs comes from donors^4^ who invest resources, largely on the basis of the funders’ goals and objectives. The paucity of local funding and underfunding of sponsored research from external funders challenges sustainability of the research enterprise in institutions/universities in LMICs^4, 5^.

Most sponsored research at IHLs is comprised of two main budgetary components; (1) direct costs and (2) indirect or overhead costs, also referred to as facilities and administrative (F&A) costs. Direct costs are costs that are directly linked to the research activities. These may include support of faculty time, graduate research assistants, post-doctoral fellows, laboratory technicians, computers and other durable equipment, and expendable commodities such as reagents, travel, communication, and clerical assistance^6^.

In HIC, F&A costs cover a portion of the IHL’s infrastructure and operational costs necessary to conduct the research, not directly linked to the research project itself. These negotiated rates are meant to cover a portion of operating, maintaining, and renovating research facilities^7^. Additionally, the indirect cost recovery supports the research enterprise by funding major research initiatives, and research and shared facilities^8^.

Although research grants cover costs directly associated with specific research projects, they do not fully cover costs that cannot be directly attributed to one project^7^. Institutions conducting sponsored research may end up spending considerable unaccounted-for amounts of funds and resources to ensure success of a project^9^. According to Dr. Kelvin K. Drogemeier, University of Oklahoma Vice President for Research, while performing sponsored research, IHLs incur a variety of other significant costs, both direct and indirect, leading up to and during a specific research project that they would otherwise not incur. Watt and Higerd^10^ and Wimsatt et al.^11^ also report that the recovered F&A costs from a grant award rarely cover the actual costs required for a research project of 24-28%^12^, and at least an additional 40% institutional costs (F&A) to cover external research funding^7^. Additionally, Association of American Universities and Colleges (AAUC)^13^ demonstrates that colleges and universities in the United States are the second-leading sponsors of research, providing about 20% of total institutional research expenditures.

According to Fonn et al.^9^ there is also the aspect of in-kind contributions (approximately 42%), that includes unremunerated labor time by faculty, use of physical and infrastructure resources, and unremunerated indirect costs in research project implementation. These contributions are provided by individual faculty who devote unremunerated time that is not counted as part of the institution’s responsibilities apart from their teaching, supervisory, and administrative roles. Without these in-kind contributions, the realization of set goals and impact of such sponsored research projects would be difficult, if not possible.

Most of the institutions in LMICs do not have policies on research costing and thus, in most instances, the F&A rates are based on funder discretion^14^. Due to this fact, there is a wide variation of indirect cost rates at IHLs in LMICs, with rates varying from 0% to 15% of direct costs^14^.

Despite an increase in funding for sponsored research in LMICs in the past decade, a quantifiable relationship between the funds requested from funding agencies and the actual cost of paying for such projects at IHLs in LMICs is poorly understood. To our knowledge, there has not been a comprehensive evaluation of adequacy of indirect costs at IHLs in LMICs to inform institutional policies on equitable distribution of research funding. Furthermore, the perceptions of stakeholders on research costing of sponsored projects at IHLs in LMICs are not well described in the current literature in order to provide direction for the initiatives to be undertaken to reduce the disparities. It is within this premise that we conducted this study to provide IHLs in LMICs the ability to quantify the actual costs of conducting sponsored research and to understand the system level factors that support or hinder the goal of “breaking even”.

## Materials and methods

This descriptive mixed methods cross-sectional study was conducted at Muhimbili University of Health and Allied Sciences (MUHAS), Kilimanjaro Christian Medical University College (KCMUCo) and Catholic University of Health and Allied Sciences (CUHAS) in Tanzania. These institutions have witnessed increased externally funded research and development project activities over the past two decades.

For the quantitative component, Audited Final Financial Reports (FFR) for externally funded research projects from January 1, 2007 and closed by December 31, 2017, were obtained from the MUHAS Office of Sponsored Projects (OSP), and the Offices of Research Management and Innovation (ORMIs) at KCMUCo and CUHAS. The FFRs contained details of specific expenditures for each project and amounts used during a specific period of funding as well as cumulative amounts for each category listed in the FFR. These categories include salaries and wages, consultancies, travel, laboratory fees, equipment, supplies and materials, training-related expenses, and regulatory fees, among others. Using the specific categories, we identified the activities undertaken to arrive at a specific expense as mandated by the institutional finance, human resources, procurement, and other departmental rules and regulations. For example, if a specific project needed to hire staff to work on the project, we worked with the human resources department and identified the cost of advertising for the position and costs associated with interviewing process hiring, Social Security contributions, payroll/wage bills, relocation/subsistence allowances and orientation/induction. These costs are expected to be paid by the IHL and not the projects because they are considered unallowable. After developing the associated costs matrix for each category, the author then applied it to both the biomedical and non-biomedical sponsored projects funded during the period previously described. For credibility, we worked closely with the respective heads of departments that perform these activities to develop the assigned associated costs from their previous experience and current costs of the activities. For biomedical projects, both equipment (depreciation, maintenance/service, clearing, insurance, bidding and disposal) and human resource components (advertising, interviewing, induction/orientation, training, relocation/subsistence, Social Security/welfare, and payroll/wage bill costs) were considered in estimating the unaccounted-for indirect costs while for non-biomedical projects the main component was human resource.

To gain insights on research costing from key stakeholders, we interviewed faculty members who had active sponsored projects or had received past sponsored project awards, senior and junior administrators and research administrators, and other junior administrators such as human resource, procurement, legal, finance, and accounting officers.

To evaluate the appropriateness of questions in the interview guide and length of the interview, six pilot in-depth interviews were conducted, and a revised interview guide was developed. The data from this pilot stage were not included in the final analysis. With purposively selected participants, face-to-face in-depth interviews were conducted by AK between March and May 2021. After providing consent, demographic information was collected and participants were asked about costing practices of sponsored projects at their institution including budgeting and F&A costs.

Statistical analysis was performed using Stata Version 15 (StataCorp LP, Texas). Descriptive statistics (median/mean and standard deviation/interquartile range) and charts were used to summarize the collected numerical data. Comparisons between biomedical and non-biomedical externally-funded research funds for unaccounted-for costs and indirect cost rates were summarized using charts.

For the qualitative component, demographic data of the in-depth interview (IDI) participants was summarized using frequency distributions. All in-depth interviews were first transcribed verbatim. Applied thematic analysis was employed to analyze segments of IDI data following a multi-stage deductive and inductive analysis approach (Kiger and Varpio 2020). Two analysts independently applied structural codes (based on the specific interview topics) to the data using Dedoose. The analysts checked for consistency by applying the code list and definitions to the transcribed interviews through discussion and reconciliation. Discrepancies in code application were resolved through discussion, and edits were subsequently made to the structural codebook, with transcripts recoded as needed. AK reviewed discrepancies between coders and acted as the tie breaker. Lastly, the analysts wrote memos to summarize the most frequently mentioned findings for each code.

Ethical clearance was obtained from the KCMUCo Ethical Review Board vide Certificate No. 2497. Permission to carry out the study was given by each respective institution’s Review Boards. Respondents were briefed about the purpose of the study and assured of strict confidentiality of the information provided and non-effect to their institutional positions.

## Results

A total of 17 projects were investigated, of which 6 (35.3%) were biomedical and 11 (64.7%) non-biomedical. Of the six biomedical donor-funded research projects, three were conducted at KCMUCo, two at MUHAS, and one at CUHAS. Of the 11 non-biomedical research projects, 2 were conducted at KCMUCo, 4 at MUHAS, and 5 at CUHAS.

The median total amount of project funding for all biomedical donor-funded research projects was United States Dollars (USD) 544,084; interquartile range (IQR) 89,268-1,226,570. The median (IQR) amount of direct costs being USD 480,600 (76,851-1,190,008) while the median (IQR) for indirect costs was USD 63,484 (12,417-117,562). The average unaccounted-for cost rate on total project cost for biomedical donor-funded research projects was 34.3% and the median (IQR) of unaccounted-for costs was USD 186,403 (30,583– 420,223). The results are shown in Table 1.

**Table 1.**
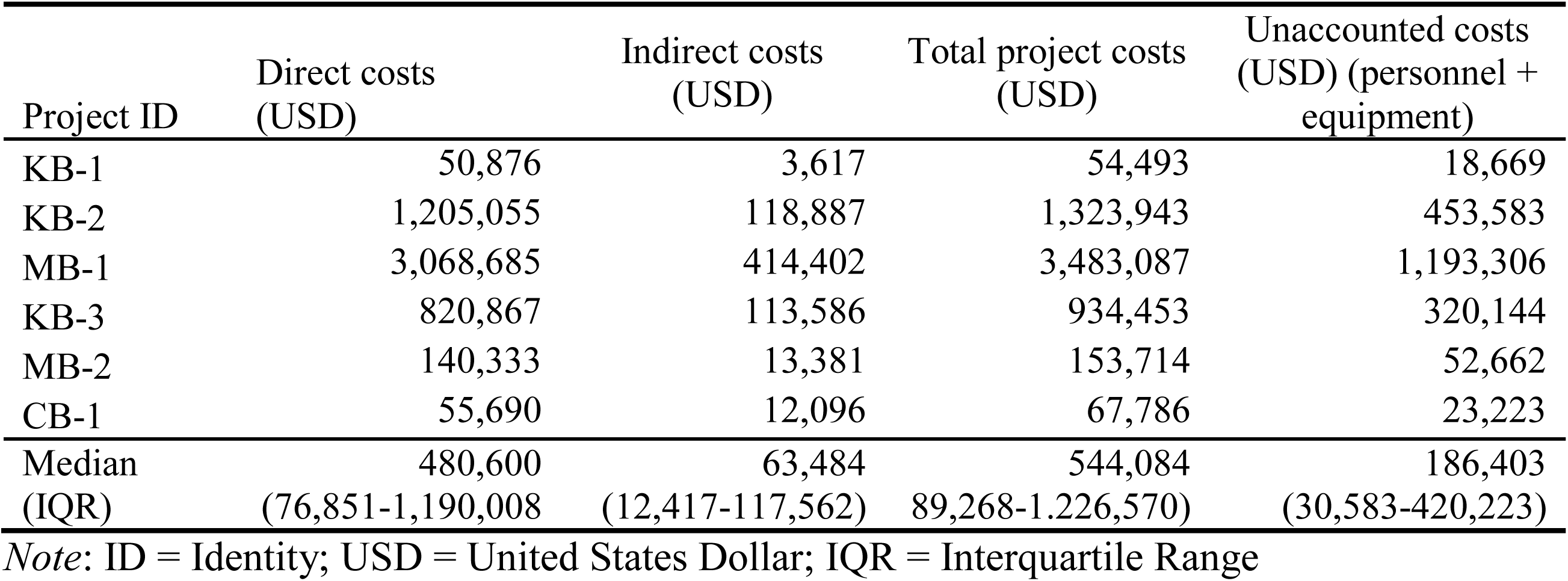
Biomedical Project Costs.

| Project ID | Direct costs<br>(USD) | Indirect costs<br>(USD) | Total project costs<br>(USD) | Unaccounted costs<br>(USD) (personnel +<br>equipment) |
| --- | --- | --- | --- | --- |
| KB-1 | 50,876 | 3,617 | 54,493 | 18,669 |
| KB-2 | 1,205,055 | 118,887 | 1,323,943 | 453,583 |
| MB-1 | 3,068,685 | 414,402 | 3,483,087 | 1,193,306 |
| KB-3 | 820,867 | 113,586 | 934,453 | 320,144 |
| MB-2 | 140,333 | 13,381 | 153,714 | 52,662 |
| CB-1 | 55,690 | 12,096 | 67,786 | 23,223 |
| Median | 480,600 | 63,484 | 544,084 | 186,403 |
| (IQR) | (76,851-1,190,008) | (12,417-117,562) | 89,268-1,226,570) | (30,583-420,223) |

*Note:* ID = Identity; USD = United States Dollar; IQR = Interquartile Range

The median (IQR) total amount of project funding for non-biomedical donor-funded research projects was USD 902,999 (468,259–1,951,212). The median (IQR) amount of direct costs was USD 790,959 (425,699–1,543,345) while the median (IQR) for indirect costs was USD 24,144 (0–137,301). In contrast to biomedical research, the median (IQR) for unaccounted-for costs (unfunded by the funder) was USD 112,875 (58,532–243,902) with the average rate on total project cost of 12.5% (Table 2).

**Table 2.**
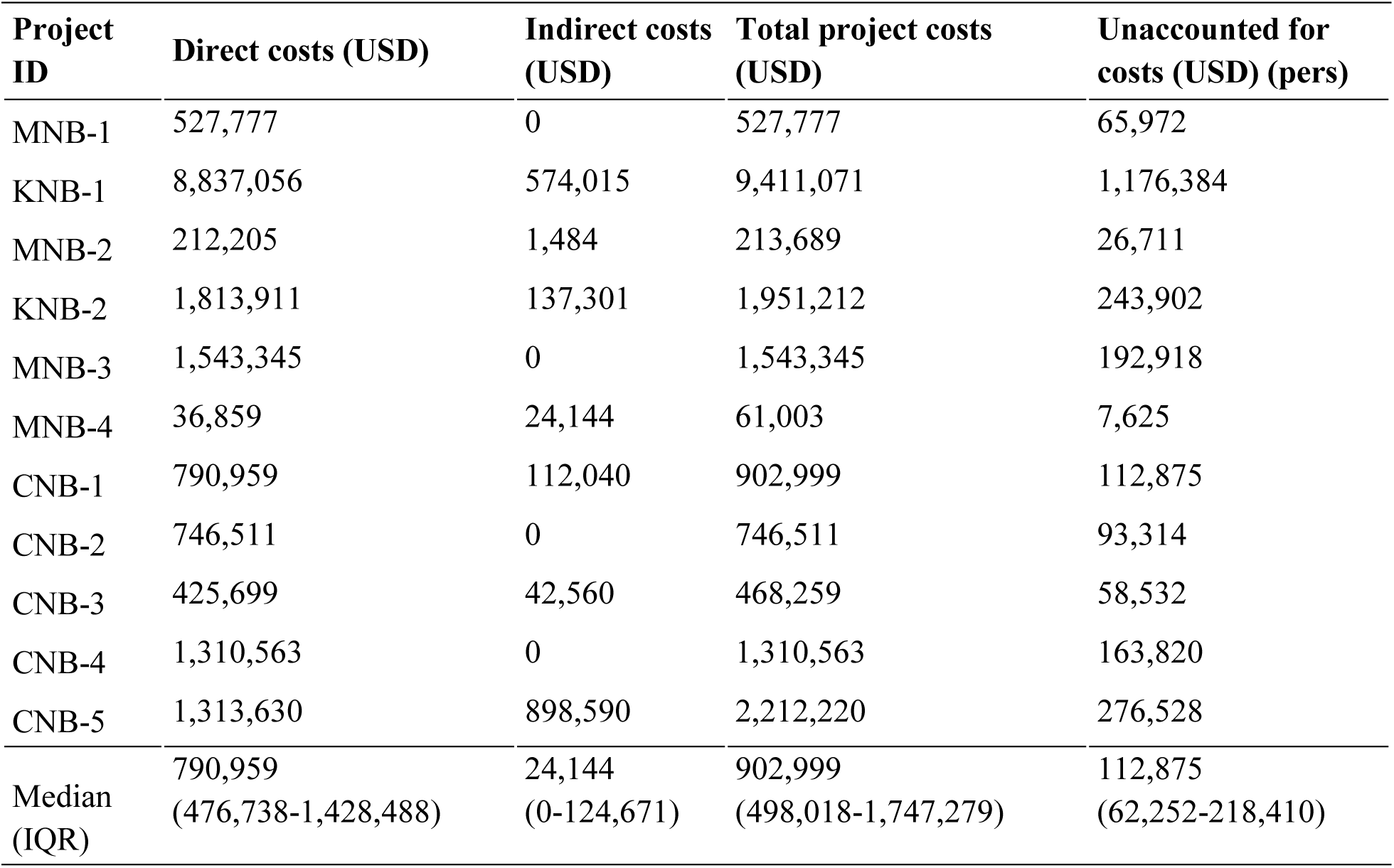
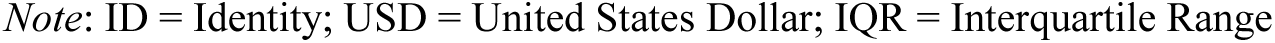
Non-Biomedical Project Costs.

| Project ID | Direct costs (USD) | Indirect costs (USD) | Total project costs (USD) | Unaccounted for costs (USD) (pers) |
| --- | --- | --- | --- | --- |
| MNB-1 | 527,777 | 0 | 527,777 | 65,972 |
| KNB-1 | 8,837,056 | 574,015 | 9,411,071 | 1,176,384 |
| MNB-2 | 212,205 | 1,484 | 213,689 | 26,711 |
| KNB-2 | 1,813,911 | 137,301 | 1,951,212 | 243,902 |
| MNB-3 | 1,543,345 | 0 | 1,543,345 | 192,918 |
| MNB-4 | 36,859 | 24,144 | 61,003 | 7,625 |
| CNB-1 | 790,959 | 112,040 | 902,999 | 112,875 |
| CNB-2 | 746,511 | 0 | 746,511 | 93,314 |
| CNB-3 | 425,699 | 42,560 | 468,259 | 58,532 |
| CNB-4 | 1,310,563 | 0 | 1,310,563 | 163,820 |
| CNB-5 | 1,313,630 | 898,590 | 2,212,220 | 276,528 |
| Median | 790,959 | 24,144 | 902,999 | 112,875 |
| (IQR) | (476,738-1,428,488) | (0-124,671) | (498,018-1,747,279) | (62,252-218,410) |

Note: ID = Identity; USD = United States Dollar; IQR = Interquartile Range

In summary, we note that (i) the larger the sponsored research fund size, the higher the unaccounted-for cost amount; and (ii) though the median total costs for non-biomedical donor-funded projects were more than 1.4 times higher than those for non-biomedical projects, the median of unaccounted-for costs for biomedical donor-funded projects was 1.7 times higher than the median of non-biomedical projects (USD 186,403 vs. 112,875,34.3 vs. 12.5% of total budgets respectively).

Figure 1 shows the comparison of the proportion of hidden/unaccounted-for costs to total direct costs for biomedical and non-biomedical sponsored research projects. It can be observed that the proportion of unaccounted-for costs for biomedical research is considerably higher than for non-biomedical sponsored research.

**Figure 1:**
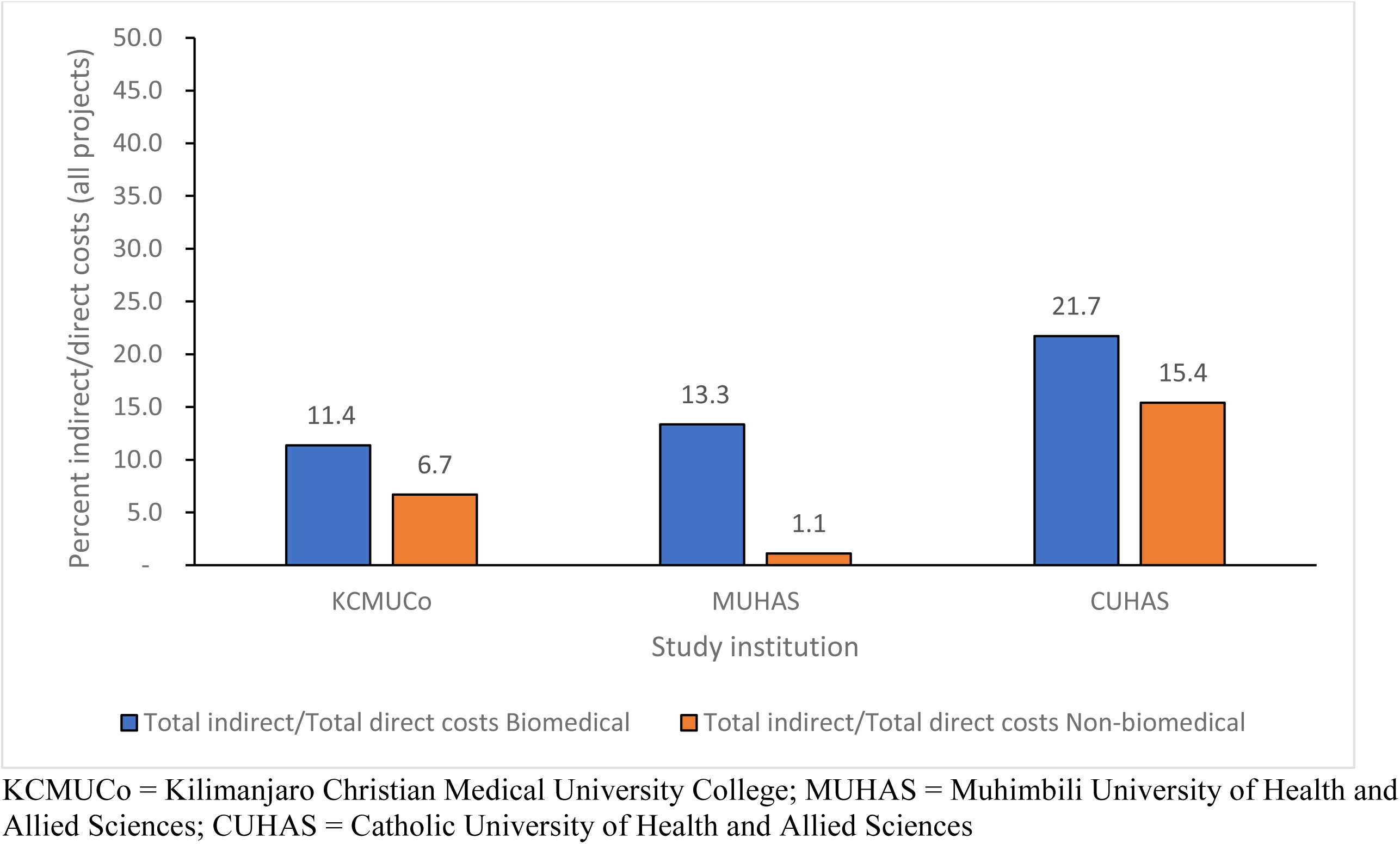
Comparison of proportion of unaccounted-for costs to total direct costs by type of project according to study institution.

Figure 2 shows the proportion of indirect costs to total direct costs, that is indirect cost rate (ICR) according to study institution by type of sponsored research (biomedical vs. non-biomedical). It can be observed that the indirect cost rates for biomedical research are relatively higher than for non-biomedical. The average indirect cost rate for biomedical research for the three study institutions is 30.9 percent (range = 13.3-57.7%) compared to non-biomedical with average ICR of 9.9 percent (range = 1.1-15.4%). In general, the average indirect cost rates for both biomedical and non-biomedical sponsored research are small.

**Figure 2:**
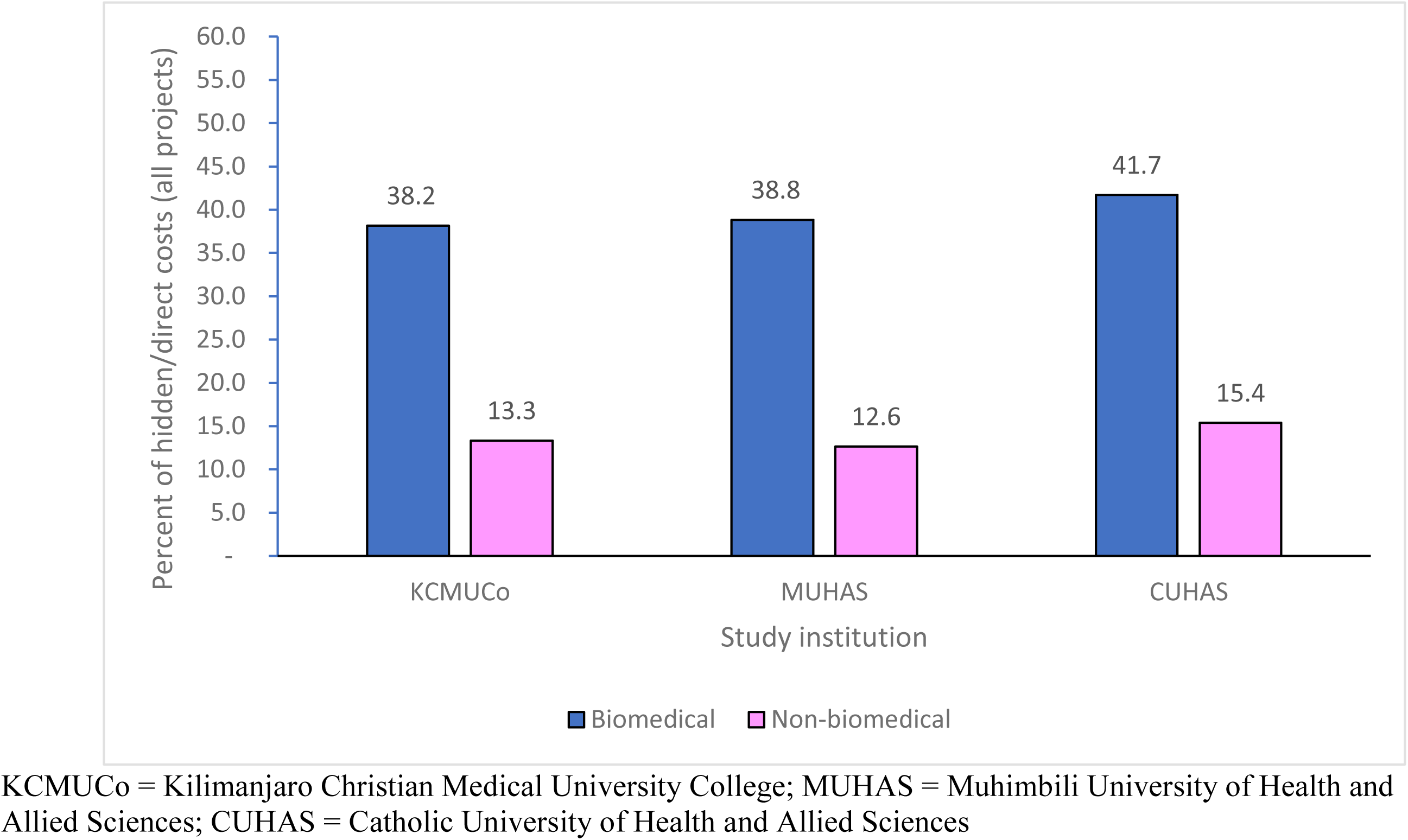
Proportion of indirect costs to total amount of direct costs by type of project according to study institution.

A total of 27 individuals participated in interviews. The majority were male, and seven held senior administrative leadership positions. Most of the participants had been with their institutions for more than 10 years. The demographic characteristics of the participants are summarized in Table 3.

**Table 3.**
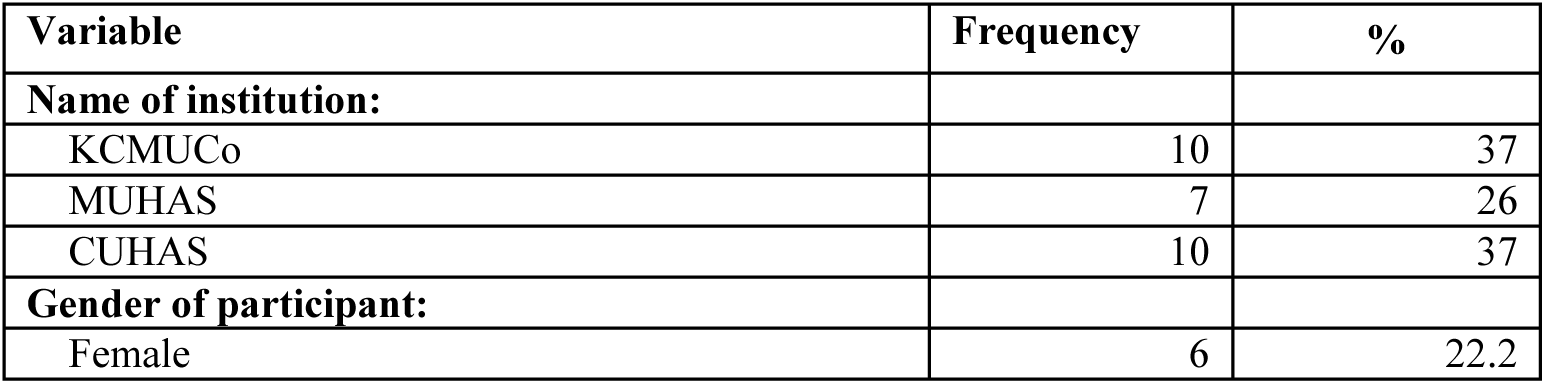
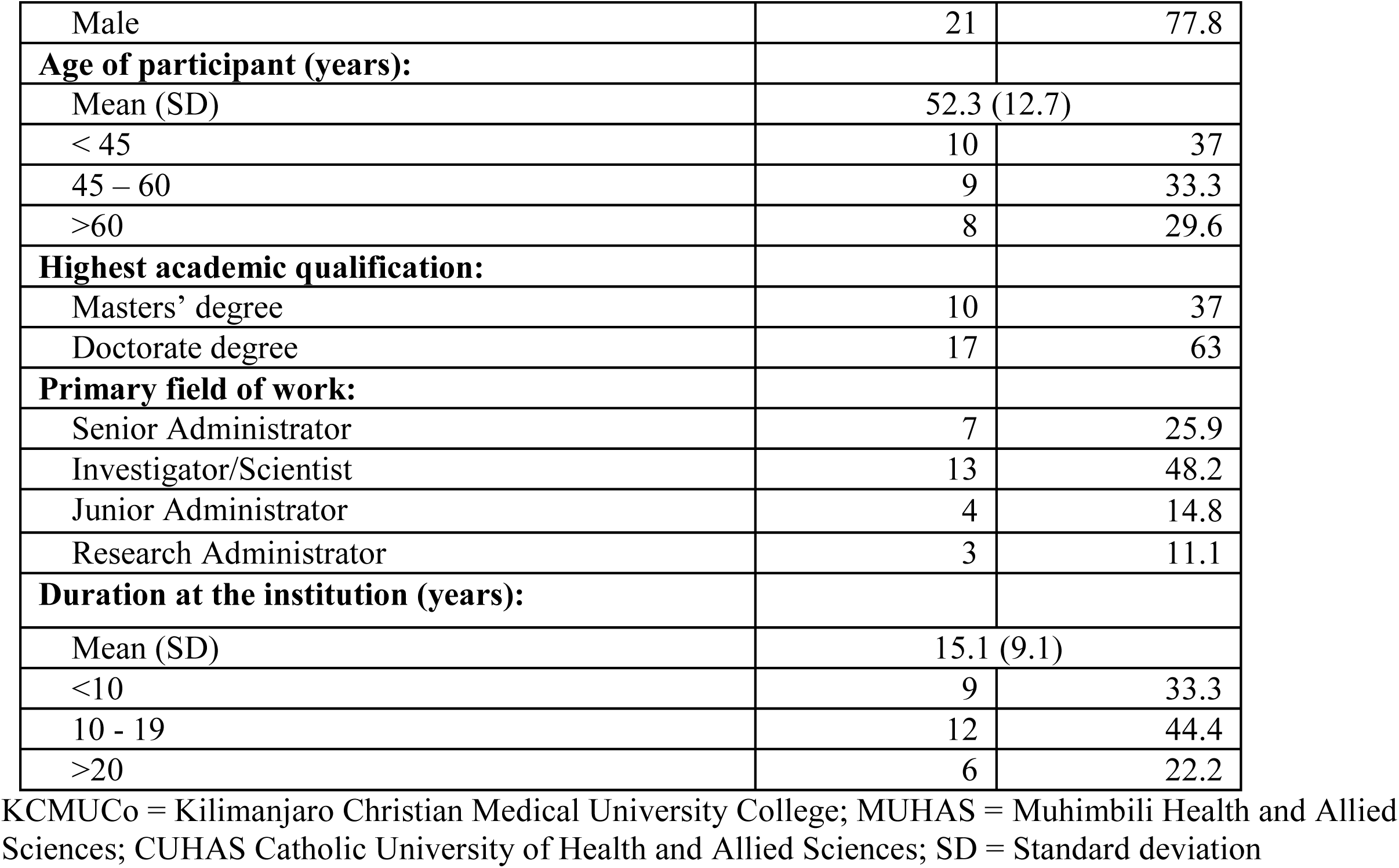
In-depth interview participants’ demographic characteristics.

Three major themes emerged from interviews with key stakeholders of sponsored research; costing practices, unaccounted-for costs, and budget negotiating power of sponsored research.

### Costing and Financial Practices Associated with Sponsored Projects

In exploring participants’ perceptions of the financial policies and procedures related to budget preparations, overwhelmingly there were issues such as determination of costs for the sponsored projects including indirect costs and their perceptions of indirect cost adequacy.

### Budgeting for Sponsored Projects

Universally, participants stated that budget development is an important step in ensuring efficient costing for research. Most participants mentioned there were no set standards used for budget development. Participants described a lack of skills in budgeting as being a limitation. Most of them highlighted the importance of training and developing the budgeting skills of researchers and administrators.

> *I think what guides budgeting is really the donor regulations on budgeting.. Some people eventually budgeted for things which are not allowable. Again, that is lack of know how in writing proposals and research administration because research administrators should be able to guide researchers. [Senior Administrator, >10, Male]*

Despite the lack of sponsored research budgeting skills, researchers were therefore expected to develop their budgets. This was not ideal, and participants recommended developing capacity in research administration.

> *Researchers cannot be experts in budgeting. You need to create that capacity for individuals, actually, even accountants who are trained in preparing budgets for research grants. Because then, they will have no conflict of interest, but they will also have a broader view of the institution to look at the visible and invisible costs. [Senior Scientist, >10, Male]*

When asked about financial policies related to sponsored projects, participants did not know of their existence at their institution. Participants reported that even though institutions had policies on research costing, there are limitations and the policies could not account for all budget and cost elements.

> *But we have policies that are guiding two things. One policy is guiding institutional overhead. Another policy is guiding the level of effort. Those are the only two things that we are worried and caring for. The rest of the things really depend on the grant. [Junior Scientist, <10, Male]*

### Indirect/Overhead and Unaccounted-for Costs

A majority of participants said that not all costs associated with a sponsored project were covered by the funding agencies. The indirect/overhead cost funds allocated by projects were not adequate to cover for unaccounted project costs. Some commented that some funding agencies had a fixed percentage for the overhead costs.

> *…..from NIH funded projects is fixed. This is supposed to cater for so many things including research administration and utilities, but they had a whole hall full of very heavy-duty freezers, you see, so you’re actually not recovering anything from the project to pay for the utilities because that overhead charge is fixed at 8%. So it was peanuts. [Senior Administrator, >10, Male]*
>
> *… don’t think so, because some of these are invisible. Are invisible costs. It’s not easy to cost them. One might argue that whatever is being provided is institution overheads, would cover some of these elements are difficult to justify in a research project. [Senior Scientist, >10, Male]*

Some participants described these standards as being ideal for local research and focused on overheads, but none existed for donor-funded research or they were not formalized.

> *We don’t have a standard, MUHAS standard, for grants. We have our institutional standards for IRB. So when you’re applying for IRB, there is a number of items that we need to hear from you, how we’re paying the staff, what will be the cost of fieldwork? What will be the cost of materials? Et cetera. Quite close to that. But we don’t have a standard or format for grants for sponsored projects, because each sponsored project, each organization has their own kind of templates, budget templates and standards. [Junior Scientist, <10, Male]*

Some of the participants lamented the lack of data and wished for a national forum to provide guidance on negotiation of overhead costs.

> *We don’t have facts and figures for negotiation. We don’t have a high policy body in the country that is speaking for all scientists in the country, that is speaking for the conduct of research in the country. You know now for example, when it comes to ethics issues, we have more voices, because in health research, it is the national health research ethics committee at NIMR which is the pillar. And our collaborators know, they require this and this and this, no squabbles. So we needed something of that nature in order to kind of come up with a formula system, and facts and figures about institutional overhead cost. Not leaving for the institutions to negotiate. [Senior Scientist, >10, Male]*

Participants also described some unaccounted costs including insurance, tax, electricity, space, water and others that were not allowable by sponsors or whose justification would require expert negotiation.

> *The other issue is insurance. It’s very uncommon to see medical equipment insured. But I think this is something that needs to move in that direction. Because people will just say, “Okay, I budget for in terms of a breakdown, I’ll do this and that.” But what about insurance?. So obviously those are things that are often overlooked in the budget, but unfortunately also often not allowed. [Junior Scientist, <10, Male]*
>
> *And it was not enough even to maintain the freezers. So that is another gray area that the imposing of a fixed overhead cost to the institutions sometimes force the institutions to pay from other sources for their utilities like water, electricity, especially electricity et cetera. Of course, there is the other thing which is not put in the equation, the people who are supporting research on a daily basis. So if your charge is only 8%, and electricity is taking more than 8%, you cannot even support the conduct of research or support research assistants who are not part of the research. [Senior Administrator, >10, Male]*

Another participant said his institution had to squeeze other operations to cover for research due to low funding.

> *You have to recast, cast down your numbers to ensure that you live within your budget. And in doing that, you find that, in most cases, you have to squeeze yourself or squeeze some operations so that you can live within your budget. That’s how we do it [Junior Administrator, <10, Male]*

### Budget Negotiating Power

A majority of the participants described the lack of negotiation power as being a big limitation in budgeting and costing for sponsored research projects. Almost all noted their institutions had a low negotiation power when deciding on rates used for budgeting.

> *Well, the budgets are developed, guided by the donor regulations that you have I think what guides budgeting is really the donor regulations on budgeting. Again, that is lack of know how in writing proposals and research administration because research administrators should be able to guide staff. [Senior Administrator, >10, Male]*

Some noted that some donor funding agencies had some room for negotiation for overheads.

> *For example, in Europe, some funding agencies were ready to negotiate, and by the time you got to negotiation, you’ve already calculated the amount of utilities’ costs and you factor that in your negotiations to make sure you’re not going to lose in terms of overhead, to make sure you’re not going to pay from your other sources to supplement utilities for research. So I very much support the idea of negotiation for overheads. [Senior Administrator, >10, Male]*

## Discussion

This study aimed to describe the costing practices of sponsored research practices in IHLs in LMICs and their implications for the unaccounted-for costs that are borne by these IHLs. Our study demonstrated that the higher the amount of the sponsored project, the higher are the unaccounted-for costs. Also, we established that despite the median total amount of non-biomedical sponsored projects being about one-and-a-half times that of biomedical projects, the median amount of unaccounted-for costs for biomedical projects was more than twice that of non-biomedical projects, which implies that unaccounted costs for biomedical sponsored research are considerably higher than those for non-biomedical sponsored research projects.

Through interviews with key stakeholders of research at three large IHLs in Tanzania, costing and financial practices associated with sponsored projects were weak, especially in budget preparation, and overhead cost estimations were inadequate. Negotiating power between funders and IHLs was also found to be lopsided, leading to unaccounted-for costs associated with sponsored projects.

The fact that the higher the total amount of the sponsored research, the higher the unaccounted-for costs could be attributed to higher indirect costs of the human resource component (for non-biomedical), including the high-tech equipment component (for biomedical projects) that are included in such projects. As expected, such projects may involve a large and diversified group of human resources for non-biomedical, and high-tech equipment for biomedical research projects, leading to increased amount of unaccounted costs.

Ehrenberg and Mykula^6^ assert that there is a perception among principal investigators that by excluding some of the indirect costs, the chance of getting funds for sponsored research increases. This propensity may be directly related to the size of the amount to be funded to the sponsored research. Furthermore, applicants may exclude some indirect costs such as salaries of administrative staff, rental of facilities, and infrastructure upgrades with the assumption that they are unallowable by the funder^14^. A US National Science Foundation report in 1991 enumerated deficiencies in calculation and application of indirect costs of federally sponsored research in educational and other institutions, including underestimating indirect costs to make an applicant’s grant more competitive and assigning responsibility for reviewing indirect costs to staff who have inadequate experience or training to accomplish the assignment effectively^15^. Also, in most instances, the indirect cost rates in LMIC are based on what the funder allows or are an estimated rate^14^. Due to this fact, there is a wide variation of indirect cost rates in LMICs, with rates varying from 0% to 15% of direct costs^14^ (ESSENCE, 2012a). As a result of these factors, the recovered indirect costs from a grant award rarely cover the actual indirect costs required for a research project^10, 11^.

This study also found that although the median amount for non-biomedical sponsored projects was higher than that of biomedical, the reverse was the case for the median amount of unaccounted costs, implying that unaccounted costs for biomedical research projects are generally higher than for non-biomedical. This could be attributed to the differences in the inherent nature of biomedical vs. non-biomedical sponsored research projects. Most biomedical research involves clinical trials, which are studies using volunteers designed to answer safety and efficacy questions about interventions^15^ and are thus characterized by longer durations, high-tech equipment, varied supplies, and larger number and more diversified human resources. Non-biomedical research in medical schools is most commonly geared toward enhancing medical education and therefore is mostly cross-sectional research primarily involving the human resource component.

Participants described weak fiscal policy pertaining to sponsored research efforts at their IHL. Given that funding agencies provide guidance or suggestions on allowable costs, budgeting policies to fully account for associated costs for sponsored research projects are important in determining the feasibility of executing a sponsored program. Negotiating power for budgeting and costing for sponsored research projects was deemed to be lopsided and favoring the funding agencies. Awards for sponsored research had a stipulated budget amount and overhead cost rate that were not negotiable. Overhead cost rates were deemed inadequate by study participants, and mostly have a fixed rate overhead rate. For example, the US NIH funds facilities and administrative costs at a fixed rate of 8 percent of modified total direct costs^16^ With this stipulation, there is need for the IHL to cover unaccounted costs as described by the participants. The continued burden of covering unaccounted-for costs at IHLs in LMICs threatens the research enterprise sustainability. It is therefore important for IHLs and funding agencies to work together to develop fair overhead rates for IHLs. Initiatives to establish offices of research and sponsored programs (ORSPs) or enhance the services of existing ORSPs that were supported by the NIH are critically needed^17^.

Results from this study should be interpreted with caution. This was an exploratory attempt to highlight costs that IHLs incur by undertaking sponsored research. First, the findings may contain inherent weaknesses pertinent to its retrospective nature such as confounding and selection bias, as well as potential unreliability of secondary data. Second, it was not possible to examine the external validity of the study for purposes of determining the generalizability of findings to other LMIC IHLs. Also, participants were recruited from three large IHLs and results may not be generalizable to all IHLs in Tanzania and other LMICs. Moreover, despite participants being reassured of anonymity of their responses with no impact on their roles at their IHL to limit social desirability bias, they may still have been inclined to respond in a positive manner. Despite the fact that the findings from this study are specific to three IHLs in Tanzania, their implications could be relevant to other medical schools and IHLs in Tanzania, sub-Saharan Africa, and LMICs worldwide. To our knowledge this is the first study that has explored the views on sponsored research of different cadres of personnel at IHLs in Tanzania.

## Conclusion

The study demonstrated that there were unaccounted-for costs for sponsored projects. We found that the larger the amount of the sponsored project budget, the higher the amount of unaccounted-for costs. This is consequential to LMIC institutions because it may be difficult to recover the full cost of a specific sponsored project, regardless of whether the project is biomedical or non-biomedical. Given this deficit in cost recovery from sponsored projects, there is a direct negative impact on their ability to sustain the research enterprise and sponsored projects may drain meager institutional resources.

Costing and financial practices associated with sponsored research projects were weak, especially in the budget preparation, and overhead cost estimation. Negotiating power for budgeting and costing for sponsored research projects was lopsided and favored the funding agencies. Awards for sponsored research had a stipulated budget amount and overhead cost rate that were not negotiable, and indirect cost rates were deemed inadequate.

It is imperative for IHLs and funding agencies to cooperate in developing equitable and fair indirect cost rates. This effort is especially important when the decolonization movement in global health is highly visible and active. In summary, there is a gap of understanding and practice of indirect cost recovery and reimbursement policies and practices within the context of sponsored research projects and the research funding model at IHLs in LMICs. IHLs and funding agencies should intentionally work to reduce the inequity in research that is currently experienced.

## Data Availability

Authors is hereby make fully available and without restriction all data underlying this this Manuscript.

## Author contributions

ATK - Conceptualization, investigation, project administration, writing the manuscript; GEK- Formal Analysis, methodology and visualization; JAB - Validation, supervision, review and editing the manuscript; CM - Methodology, data curation and original draft preparation; and SdeJ – Supervision

## Acknowledgements

The authors thank all the stakeholders including study participants, Leadership at MUHAS, KCMUCo, CUHAS who participated to provide inputs to develop this piece of work that advocates resources’ equity across the globe.

